# Characterization of an Arctic-like 1a rabies virus from a 54-day-old puppy with atypical presentation, Pune, India, 2026

**DOI:** 10.64898/2026.07.09.26357633

**Authors:** Padinjaremattathil Thankappan Ullas, Vikas Sharma, Veena Vipat, Shweta Choudhary, Alfia Fathima Ashraf, Ranjana Mariyam Raju, Venu Kotturi, Kunal Sudhakar Sakhare, Vijay Parashramji Bondre

## Abstract

Rabies remains a significantly underreported fatal zoonosis in India, where the Arctic-like 1a (AL1a) lineage predominates in dog populations. While atypical clinical presentations in dogs can delay diagnosis and increase human exposure risk, genomic and clinical data on neonatal canine rabies remain limited. This study reports an exceptional case of rabies in a 54-day old unvaccinated German shepherd puppy which presented with severe pruritus and self-biting behaviour. The puppy was euthanized due to poor clinical response. Post-mortem testing revealed viral antigen (by Direct Fluorescent Antibody Test) and viral RNA (by real-time RTPCR) in the brain tissue. Whole-genome sequencing recovered a near-complete rabies virus genome (11,947 nucleotides; 99.5% genome coverage), classified within the AL1a_A1.1 sublineage. Phylogenetic analysis revealed close genetic relatedness to contemporary Indian rabies virus strains. Comparative genomic analysis identified 4, 3, 6, and 8 non-synonymous substitutions in the phosphoprotein, matrix, glycoprotein, and polymerase genes, respectively. This case is one of the youngest documented cases of canine rabies with atypical manifestations, caused by the AL1a viral clade. Our findings highlight the risks associated with neonatal canine rabies, the need for heightened clinical suspicion in atypical cases, and the importance of genomic surveillance to monitor evolving rabies virus lineages in endemic regions.

## Introduction

Rabies is a highly underreported fatal zoonotic encephalomyelitis, caused by RNA viruses of the Genus *Lyssavirus* within *Rhabdoviridae.* Rabies claims around 59,000 human lives and thousands of livestock globally every year (1), though death estimates from global rabies burden studies range from 20,000 to 175,000 (2). Dog-mediated human rabies deaths persist in more than 100 countries (2). While high-income countries have nearly eliminated canine rabies, the disease remains endemic in developing countries in Asia and Africa (3). India remains a global rabies hotspot, reporting about 36% of the global human rabies deaths (1). Rabid dogs are responsible for more than 99% cases of disease transmission. Though India rolled out a comprehensive National Action Plan for Dog-Mediated Rabies Elimination, major gaps remain in the surveillance of canine rabies and effective animal birth control.

The Genus *Lyssavirus* currently includes 18 named species, including *Lyssavirus rabies*, the causative agent of classical rabies (ICTV). The canine/terrestrial carnivore-related lyssaviruses are classified into 6 major global clades: Cosmopolitan, Arctic & Arctic-like, Asian, Indian Subcontinent, Africa-2, and Africa-3 (4). Two distinct lineages of rabies virus (RABV), viz., the Arctic-like (1a and 1b sublineages) and the Subcontinental lineage are prevalent in India. The majority of the Indian isolates belong to the Arctic-like 1a sublineage, though strains from north and south India show distinct clustering. A recent study also identified a distinct north-eastern clade within the AL1a lineage (5).

Standard veterinary guidelines recommend rabies vaccination of dogs at 12 weeks of age (considering persistence of maternally derived antibodies) and a subsequent booster (6). However, this age gap leaves a period of high vulnerability for very young puppies born to unvaccinated free-roaming dogs or with inadequate transfer of maternal antibodies. This also creates a public health blind spot, involving low risk perception for rabies in very young puppies, and neglect of behavioural and atypical symptoms of rabies in them. Higher incidence of rabies has been reported in dogs younger than 12 months, including puppies under 3 months of age, in rabies-endemic countries (7).

While rabies in canines usually presents either in the ‘furious’ (encephalitic) form characterized by aggression and behavioural symptoms or the ‘dumb’ (paralytic) form marked by paralysis, atypical presentations have also been recognized. The authors have recently identified an atypical canine case of rabies in an adult dog which presented with aggravation of epilepsy. RABV replication within dorsal root ganglia can result in localized or generalized intense pruritus and is frequently seen in patients with rabies (8).

Despite the well-documented epidemiology of Arctic-like 1a strains in India, comprehensive genomic characterization attempts of such strains associated with neonatal canine rabies have been rare. We report an exceptional case of an unvaccinated puppy, which exhibited atypical manifestations including severe itching, and self-biting behaviour necessitating euthanasia, in which rabies due to an Arctic-like 1a strain was confirmed by molecular testing.

### Case presentation

A 50-day-old male German shepherd puppy was brought by its owners to a local veterinary clinic in Pune, Maharashtra on 3^rd^ April, 2026, with complaints of severe itching on the lower abdomen and a scabies-like patch on the right ear. The puppy was dewormed with oral fenbendazole (one dose each on 24^th^, 25^th^ and 26^th^ April), and vaccinated against canine parvovirus and distemper (Puppy DP) on 27^th^ March 2026. On 4^th^ April, the puppy presented with complaints of severe itching of the lower back and inguinal area, and was treated with intramuscular ceftriaxone and prednisolone followed by oral amoxicillin-clavulanic acid (0.9 mL bd) and gabapentin (25 mg, hs) starting the next day. On 5^th^ April, the puppy again presented with intense itching and self-biting. On examination, there was constant itching on the right ear, belly and side of the trunk. The pupillary light reflex was normal, and there was no nystagmus. Fleas were noted around the head. The puppy was treated with oral cetirizine (4 mL od) and enrofloxacin-silver sulphadiazine ear drops (o.d. and s.o.s.). On 7^th^ April, the puppy had one episode of vomiting, persistent itching of the right ear, biting of the right thigh, and was vocalizing constantly. Hematological tests revealed leucocytosis (29,000 cells/mm^3^), lymphopenia (4.5%), neutrophilia (90.9%), and thrombocytosis (574,000 cells/mm^3^). An X-ray of the skull was unremarkable. Biochemical testing revealed serum creatinine of 0.24 mg/dL, serum glutamic pyruvic transaminase of 41 U/L, serum sodium of 144 mEq/L, serum potassium of 4.3 mEq/L, and serum chloride of 113 mEq/L. The puppy was treated with oral ranitidine (0.3 mL t.d.s.), augmentin (1.1 mL b.d.), Relaxzyme-S (1 o.d.), gabapentin (15 mg, o.d.), ondansetron (3.1 mL b.d.) and Comfoderm spray (anti-inflammatory). By 8^th^ April, the puppy had 3 episodes of vomiting. On examination, the puppy was hyperactive, biting aimlessly, vocalizing constantly, and febrile (initially 103°F, then 105°F). Rapid lateral flow antigen tests for canine parvovirus and rabies performed at the clinic yielded negative results. On 9^th^ April, the puppy was active but mostly recumbent, had two episodes of vomiting, and exhibited microseizures and chorea. The puppy was started on intravenous fluids, and treated with intravenous pantoprazole (40 mg), ceftriaxone (1000 mg), ondansetron (30 mL), oral phenobarbital (0.6 mL bd) and oral gabapentine (50 mg, 1 od). Lateral flow rapid antigen test for canine distemper virus yielded a negative result. During the period of sickness, three family members suffered bites from the puppy and sought post-exposure rabies prophylaxis. In view of poor clinical outcome, the puppy was euthanized on 9^th^ April, 2026.

## Methods

Post-mortem brain tissue of the puppy was transported to the Rabies Laboratory under cold chain conditions and processed immediately.

### Direct Fluorescent Antibody Test for rabies

Impression smears were prepared from the tissue, and a Direct Fluorescent Antibody Test was performed using a FITC anti-rabies monoclonal globulin (Fujirebio, Inc.) as per manufacturer instructions.

### Real-time PCR for rabies

Total RNA was extracted from 100 mg of the tissue specimen using TRIzol^TM^ reagent (Thermo Scientific) as per manufacturer instructions. TaqMan real-time reverse transcription polymerase chain reaction (RTPCR) for rabies virus was performed using a standard protocol (9). The assay was performed using 5 µL of total RNA extract from brain tissue and AgPath-ID™ One-Step RT-PCR reagents (Thermo Scientific) on a Bio-Rad CFX96 real-time PCR detection system.

### Next Generation Sequencing

RNA extracted from the brain tissue specimen was quantified using the Qubit RNA HS Assay Kit (Thermo Fisher Scientific, Waltham, MA, USA). A total of 100 ng of RNA from the sample was used for library preparation with the Illumina RNA Prep with Enrichment (L) Tagmentation Kit (Cat. No. 20040536; Illumina, Inc., San Diego, CA, USA), which incorporates first- and second-strand cDNA synthesis, tagmentation-based fragmentation, adapter/barcode ligation, and library amplification. Target enrichment was performed using the Illumina Virus Surveillance Panel V2 (Cat. No. 20108081), employing hybrid-capture enrichment probes for approximately 200 viral pathogens. The pooled, enriched library was normalized to 10 pM and sequenced on the Illumina MiSeq platform using a MiSeq Reagent Kit v3 (150 cycles; Cat. No. MS-102-3001), generating a total of 0.89 million paired-end reads with a mean target coverage depth of 3368×. Raw FASTQ files (R1 and R2 reads) were processed using the DRAGEN Microbial Enrichment analysis tool (Version 4.5.4000) on the Illumina BaseSpace Sequencing Hub using default parameters. Sequence data have been deposited in the NCBI Sequence Read Archive under accession number [submission in progress].

### Bioinformatics Analysis

#### Assembly and Annotation

Raw paired-end Illumina sequencing reads (2 × 76 bp) were assessed using FastQC (v0.12.1)(10) and quality filtered using Fastp (v1.3.3)(11). This process included adapter trimming, a minimum Phred quality score of 20, and a minimum post-trimming read length of 50 bp. The filtered reads were aligned to the Arctic-Like 1a reference genome (GenBank accession PP737617; highest-quality complete genome from the same lineage) using BWA-MEM (v0.7.17)(12) with default parameters. The alignments were sorted and indexed using SAMtools (v1.20)(13), and variants were called using BCFtools mpileup (v1.20; maximum depth = 1000) followed by BCFtools call (14). Consensus genome sequences were generated using <u>vcf2consensus.pl</u> (https://github.com/ellisrichardj/csu_scripts/tree/master) (15) with a minimum depth threshold of 20X; positions below this threshold were masked as ‘N’. Genome coverage, breadth of coverage, and sequencing depth statistics were calculated from BAM alignment files using SAMtools *coverage* and *depth* commands. Read mapping, variant calling, consensus sequence generation, and assembly quality assessment were performed through a custom workflow incorporating the tools described above. Lineage assignment of the novel sequence was performed using RABV-GLUE (16) and MADDOG lineage-typing tool (17).

#### Sequence Retrieval

For phylogeographic analysis, a curated subset of RABV genomes was constructed based on established lineage references. The initial dataset comprised 624 genomes in total, including 145 representative Indian and regional sequences, 44 global lineage reference and outgroup sequences (Arctic AL1a–AL3, AL2, Indian-Subcontinent, Cosmopolitan AF1a–AF4, and Asian SEA1–2 lineages) retrieved from the RABV-GLUE database, and the novel genome generated in this study. To reduce redundancy, highly similar genomes were clustered using CD-HIT (cd-hit-est) v4.8.1 (18) with a 99.5% nucleotide identity threshold. One representative sequence per cluster was retained, yielding a non-redundant dataset of 86 genomes for downstream analysis.

#### Multiple Sequence Alignment

All 86 RABV genomes were aligned at the complete-genome level using MAFFT (v7.525) (19) with the --auto strategy to automatically select the optimal algorithm. The resulting alignment was trimmed using ClipKIT (v1.4.2) (20) in smart-gap mode to remove poorly aligned and highly gapped positions while retaining phylogenetically informative sites.

#### Phylogenetic Reconstruction

Maximum-likelihood phylogenetic analysis of the 86 genome dataset was executed using IQ-TREE 2 (v2.2.6, MPI version) (21). The best-fit nucleotide substitution model (GTR+F+R6) was selected using ModelFinder; branch support was assessed using 1,000 ultrafast bootstrap replicates and used for maximum-likelihood tree reconstruction.

#### t-SNE Clustering

To complement phylogenetic inference and visualize genome-wide genetic relationships among rabies virus (RABV) isolates, t-distributed stochastic neighbor embedding (t-SNE) was applied to a dataset of 600 complete, high-quality RABV reference genomes, independent of the phylogenetic dataset. Pairwise genomic distances were first computed from the whole-genome alignment and used as input for dimensionality reduction using the **scikit-learn** package (v1.3.0) in Python. The optimal number of clusters was determined using the elbow method, followed by K-means clustering. Dimensionality reduction and two-dimensional visualization were conducted using the scikit-learn package in Python (v1.3.0) (22).

#### Mutation Analysis and Structural Alignment

Genomic variations in the novel study sequence were characterized through comparative mutational analysis using an in-house Python script against the AL1a reference genome (GenBank: KF154996). To evaluate the biological implications of non-synonymous substitutions, 3D structural alignments were performed between predicted protein models of the isolate and corresponding wild-type structures using the Protein Structure Alignment workflow in the Schrödinger suite.

#### Selection Pressure Analysis

The coding regions (CDS) of all five RABV proteins (G, M, N, P, and L) were analyzed for selection pressure. Pervasive purifying and positive selection were determined using the Single-Likelihood Ancestor Counting (SLAC) approach (23). The Mixed Effects Model of Evolution (MEME) (24) was used to detect sites under positive (diversifying) selection. Both analyses were conducted using the standard viral genetic code (translation table 11) with default settings and a significance threshold of p ≤ 0.1 implemented in HyPhy through the Datamonkey web server (25).

## Results

### Detection of rabies virus antigen

Direct Fluorescent Antibody Test performed on acetone-fixed impression smears prepared from the brain tissue specimen revealed strong positivity for rabies virus nucleoprotein antigen (Fig. 1), confirming rabies encephalitis.

**Fig 1.**
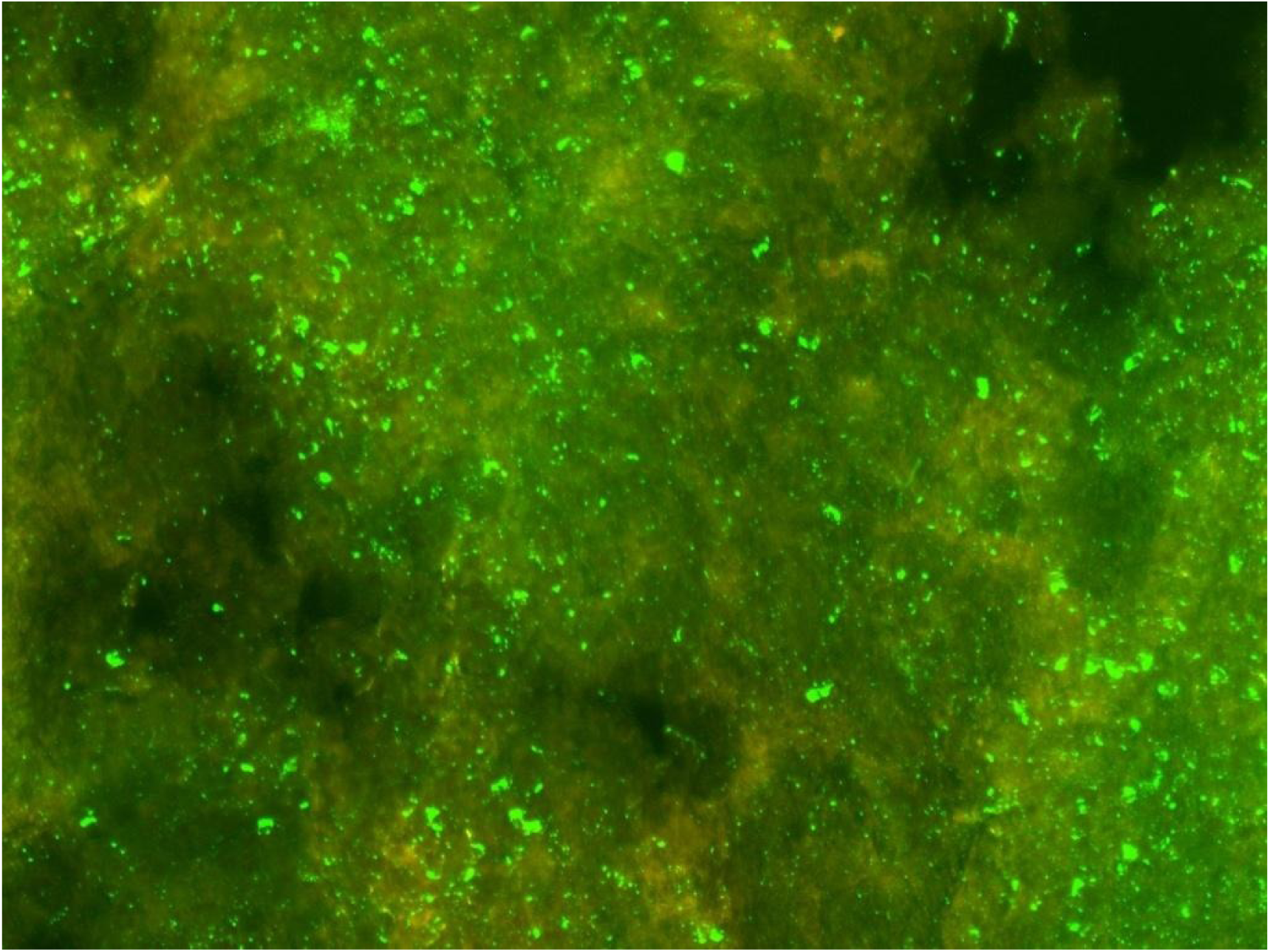
Direct Fluorescent Antibody Test on the brain tissue specimen, showing positivity for rabies virus nucleoprotein antigen (punctate to diffuse bright apple-green fluorescence within cytoplasm and neuronal processes) (420X).

### Detection of rabies virus RNA

Real-time RT-PCR on total RNA extracted from brain tissue detected rabies virus RNA (mean cycle threshold (Ct): 25.63 from duplicate wells). Successful co-amplification of the host beta-actin control confirmed sample quality and RNA integrity.

### High-Quality Reference-Based Genome Assembly and Lineage Annotation

Reference-guided assembly produced a complete genome sequence of 11,788 bp. The assembled genome exhibited a GC content of 45.55% and contained no ambiguous nucleotides. The consensus sequence achieved 100% genome coverage with a mean sequencing depth of 3,842.67X. Coverage was uniformly distributed across the genome, with depths ranging from 43X to 9,187X. All genomic positions were covered at depths ≥10×, while 99.96% and 99.90% of positions were covered at depths ≥50× and ≥100×, respectively, indicating a high-confidence assembly suitable for downstream genomic and phylogenetic analyses. The analysis using RABV-GLUE identified the virus as belonging to the Arctic-like lineage 1a and further characterized it as the sub-lineage AL1a_A1.1. Independent verification using MADDOG gave the same classification. According to the metadata, the AL1a_A1.1 sub-lineage was found only in India between 2003 and 2023.

### Phylogenetic Analysis

For phylogenetic reconstruction, a curated, non-redundant dataset was generated by clustering sequences with CD-HIT, yielding 86 representative genomes for tree inference. The multiple sequence alignment was then trimmed with ClipKIT, which removed 52 poorly aligned or gapped positions and reduced the alignment length from 11,947 to 11,895 sites (0.44% of total positions), while retaining the vast majority of informative sites. Maximum-likelihood analysis resolved the major global rabies virus (RABV) lineages into distinct, well-supported clades, with most internal nodes receiving strong ultrafast bootstrap support (≥70) (Fig. 2). The phylogenetic topology was consistent with the established classification of Arctic, Indian Subcontinent, Cosmopolitan, and Asian lineages. The Arctic AL1a lineage formed a monophyletic clade comprising isolates predominantly from northern and northeastern India and neighboring regions. Within this lineage, sequences generally clustered by geographic origin, with relatively short branch lengths indicating limited genetic divergence among regional isolates.

**Fig 2.**
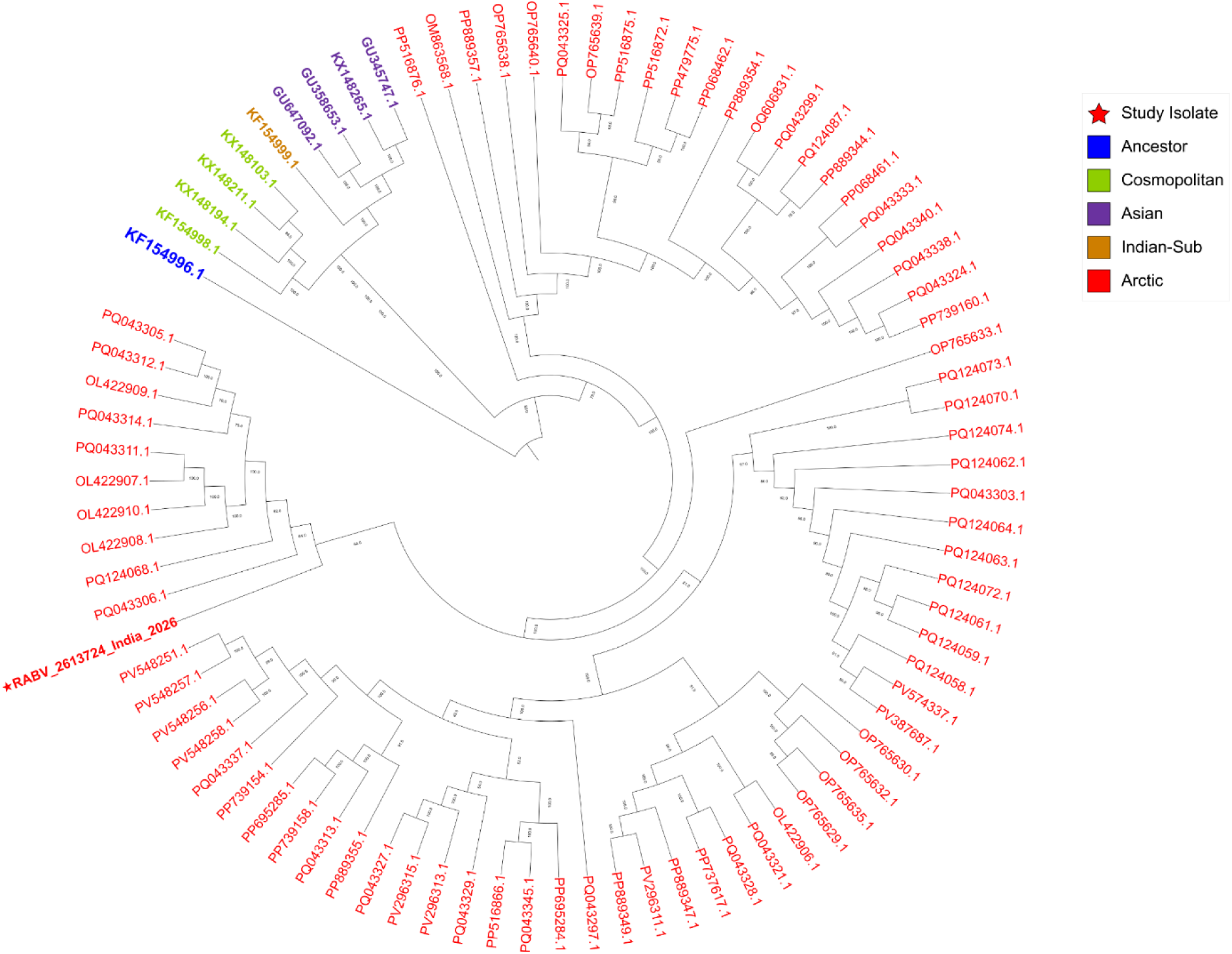
Whole-genome phylogenetic tree of rabies virus. Phylogenetic tree constructed by analyzing whole-genome nucleotide sequences. Colored circles denote the different clades identified in the phylogenetic analysis; the star indicates the isolate characterized in this study; and the black square represents the ancestral reference sequence.

The isolate characterized in this study (RABV_2613724_NIV_India_2026) clustered within the Arctic AL1a lineage and showed close relatedness to previously reported AL1a isolates from India. Its short branch length and strong bootstrap-supported placement indicate a high degree of genomic similarity to contemporary regional AL1a viruses, supporting its assignment to the Arctic AL1a lineage (Fig. 2).

### t-SNE Clustering

The results from the t-SNE analysis showed distinct clusters representing global lineages, including Cosmopolitan, Asian, Indian Subcontinent, and Arctic clades. The observed clustering pattern was consistent with the maximum-likelihood phylogeny (Fig. 2), with genomes belonging to the same phylogenetic lineage occupying discrete regions of the two-dimensional embedding.

The study isolate clustered within the Arctic AL1a group and closely overlapped with contemporary Indian AL1a genomes, indicating high genome-wide similarity to locally circulating Arctic AL1a viruses.

**Fig 3:**
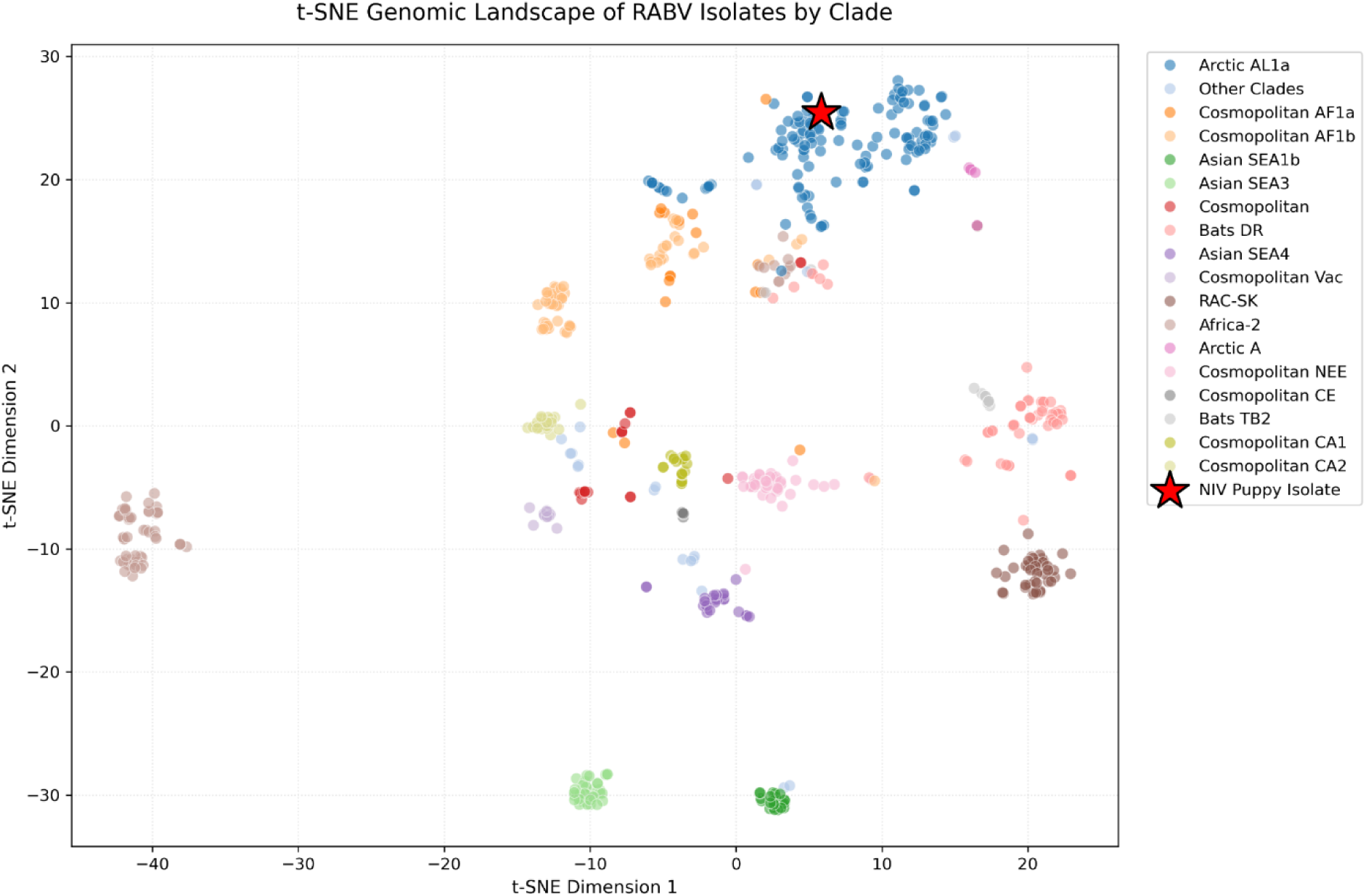
t-SNE plot of RABV genomes by clades. t-SNE visualization of global rabies virus genomes by clade. Each point represents a single RABV genome projected into two-dimensional space based on whole-genome sequence similarity. Points are colored by major lineage (Cosmopolitan, Asian, Indian Subcontinent, Arctic and Arctic-like, and other reference clades), forming distinct, well-separated clusters that reflect known phylogenetic groupings. The study isolate is embedded within the Arctic-like cluster, closely overlapping with contemporary Indian AL1a sequences, indicating strong genomic similarity to locally circulating strains and no evidence of drift toward other global lineages.

### Mutational Landscape and Structural Preservation

When the Indian sequence was compared against the AL1a rabies reference genome (KF154996), 21 non-synonymous mutations were identified across the viral genome: L (n=8), G (n=6), M (n=3), P (n=4), and N (n=0). The notable substitutions found in the Polymerase are E18D, A19V, P24A, R48K, and G2097E, alongside localized changes in the Phosphoprotein (Table 1). The Structural superimposition of the modeled isolate against the wild-type reference yielded a highly concordant Alignment Score of 0.024 and a significantly low Root Mean Square Deviation (RMSD) of 0.568 Å. The low RMSD (0.568 Å) indicates a high degree of structural similarity between the predicted isolate model and the reference structure, suggesting that the overall protein fold is largely preserved despite the observed amino acid substitutions.

**Table 1:** List of mutations observed in the isolate relative to reference sequence KF154996.

| S. No. | Gene | Genomic Coordinate | Nucleotide Change | Amino Acid Change | Amino acid Position in Region | HGVS Notation |
| --- | --- | --- | --- | --- | --- | --- |
| 1 | L | 54 | gag → gat | E → D | 18 | E18D |
| 2 |  | 56 | gcg → gtt | A → V | 19 | A19V |
| 3 |  | 70 | ccc → gcc | P → A | 24 | P24A |
| 4 |  | 143 | aga → aaa | R → K | 48 | R48K |
| 5 |  | 3406 | acg → gca | T → A | 1136 | T1136A |
| 6 |  | 4681 | agt → ggt | S → G | 1561 | S1561G |
| 7 |  | 4855 | gat → aat | D → N | 1619 | D1619N |
| 8 |  | 6290 | gga → gag | G → E | 2097 | G2097E |
| 1 | G | 209 | acc→ atc | T→I | 70 | T70I |
| 2 |  | 1124 | aag→agc | K→S | 375 | K375S |
| 3 |  | 1373 | agg →aag | R→K | 458 | R458K |
| 4 |  | 1423 | ctc→ttc | L→F | 475 | L475F |
| 5 |  | 1431 | ata→atg | I→M | 477 | I477M |
| 6 |  | 1516 | agc→ggc | S→G | 506 | S506G |
| 1 | M | 133 | aca→gca | T→A | 45 | T45A |
| 2 |  | 170 | gaa→gga | E→G | 57 | E57G |
| 3 |  | 230 | aga→aaa | R→K | 77 | R77K |
| 1 | P | 194 | ggc→gcc | G→A | 65 | G65A |
| 2 |  | 388 | gcg→aca | A→T | 130 | A130T |
| 3 |  | 485 | tcc→tac | S→Y | 162 | S162Y |
| 4 |  | 521 | gtg→gcg | V→A | 174 | V174A |

### Selection Pressure

Codon-based analysis utilizing the SLAC method revealed extensive negative selection constraints across the viral genome, identifying 276 of 524 sites in the glycoprotein, 266 of 450 sites in the nucleoprotein, 107 of 202 sites in the matrix protein, 151 of 297 sites in the phosphoprotein, and 1197 of 2127 sites in the polymerase. In contrast, evaluation with MEME detected diversifying selection within the matrix protein (codons 77 and 151), glycoprotein (codons 169, 478, 484, and 492), phosphoprotein (codon 162), and polymerase (18 discrete sites, notably including positions 19, 429, 434, 435, 437, 438, 442, 443, 847, and 1050).

## Discussion

This study provides a comprehensive genomic, structural, and evolutionary characterization of a novel Indian rabies virus strain (RABV_2613724_NIV_India_2026) associated with an atypical case of canine rabies. The strain was detected in a 54-day-old unvaccinated puppy, representing a remarkably young age for canine rabies infection. Prior reports have indicated that rabies may occur in very young puppies, posing a significant public health problem (26) (27). Unusual symptoms such as severe pruritus and self-biting behavior, as in the present case could easily be misinterpreted for dermatological, neurological, or behavioral disorders, delaying early diagnosis and increasing exposure risks to handlers. This is particularly important in rabies-endemic regions, where early clinical symptoms directly influence post-exposure prophylaxis decisions and public health response. Confirmation of rabies diagnosis by complementary approaches such as viral antigen detection by the fluorescent antibody test, viral RNA detection (by real-time RT-PCR) and whole genome sequencing (where feasible) can help to avoid delayed or missed diagnosis in cases with atypical presentations. Sequencing-based surveillance, by deciphering the evolutionary context of circulating virus strains and tracking lineage persistence, can add further value to rabies diagnosis, and strengthen routine rabies control programs, especially in rabies-endemic regions (28).

By positioning the full genome of the isolate within a curated, non-redundant dataset of 86 global and regional reference sequences, we inferred the evolutionary dynamics and structural adaptations essential to this endemic viral population. In accordance with established epidemiological records, lineage assignment tools classified our isolate as belonging to the Arctic-like 1a (AL1a) lineage, specifically the AL1a_A1.1 sub-lineage. The study isolate grouped closely with contemporaneously circulating regional strains, exhibiting remarkably low genetic divergence. This is consistent with observations from a recent comprehensive study of Indian RABV strains by Varun et al. (2026) (5). The localized phylogeographic pattern highlights the role of constrained viral dispersal. The tight phylogenetic grouping and highly conserved structural topology suggest that the virus is deeply integrated into the stable genetic framework of the regional endemic population, reflecting continuous local transmission rather than a distant introduction.

This localized endemicity is further supported by our high-dimensional t-SNE clustering analysis of 600 global sequences. The spatial coordinates clearly delineated major global clades such as the Cosmopolitan, Asian, and Indian Subcontinent lineages, and demonstrated that our study sequence clearly groups within the Arctic lineage cluster. This clustering pattern is consistent with the phylogenetic analysis and further supports the assignment of the isolate to the Arctic AL1a lineage..

To understand how this sustained endemic circulation influences the virus, we evaluated its mutational and structural landscapes. Comparative sequence analysis against the AL1a reference genome identified several non-synonymous mutations in the G and L genes, highlighting ongoing adaptation dynamics or neutral drift during endemic circulation, as these surface (G) and replicative (L) proteins generally exhibit greater tolerance in sequence variability than internal components (29). The high degree of conservation of nucleoprotein is consistent with its recognition as the most stable and structurally constrained gene among lyssaviruses (30). 3D structural superimpositions of the isolate’s modelled proteins onto wild-type references revealed highly concordant alignments with sub-Angstrom RMSDs, indicating conservation of the overall 3D backbone folding and essential structural architecture of the viral replicative machinery (31).

The conserved structural features also indicate persistent evolutionary pressure. Codon-based SLAC analysis revealed pervasive purifying selection across the genome [a phenomenon well documented in large-scale lyssavirus phylogenomics datasets (4)], reflecting a strict biological requirement to conserve core viral functions. Despite the high degree of conservation, MEME analysis identified discrete loci with diversifying selection in the polymerase, glycoprotein, and phosphoprotein genes, indicating preferred targets for episodic adaptation during endemic maintenance and host-shift barriers (4). The findings indicate that the regional RABV population preserves a rigid physical outline through extensive negative selection while simultaneously introducing site-specific mutations to enhance viral fitness and local adaptation.

## Conclusion

Overall, this isolate highlights that rabies remains a persistent endemic threat within canine populations in India, driven by the uninterrupted local circulation of AL1a viruses. Rabies should be suspected and investigated in any unvaccinated puppy exhibiting abnormal behavior or neurological signs, regardless of age, and genomic surveillance should augment conventional diagnostics wherever feasible. This case underscores the need to re-examine regional rabies vaccination policies in puppies - particularly the age of first immunization in endemic settings. In low- and middle-income countries (LMICs), unregulated commercial companion animal sales present a severe public health hazard, as puppies are frequently traded well before reaching the standard three-month rabies vaccination threshold. Mitigating this risk requires robust municipal health policies that strictly enforce breeder licensing, pet-sale tracking, and mandatory point-of-sale veterinary health certification. Combining these regulatory frameworks with targeted public awareness campaigns that empower consumers to demand maternal immunization records will be critical to closing this urban transmission loophole and achieving sustainable rabies elimination. Collectively, these findings demonstrate the value of integrating whole-genome sequencing with phylogenetic, structural, and evolutionary analyses to improve rabies surveillance and inform evidence-based control strategies in endemic regions.

## Data Availability

The whole-genome sequence generated in this study has been deposited in GenBank under accession number PZ638805. All other data supporting the findings of this study are included within the article and its supplementary information files.

## Author contributions

PTU and VS conceived the study; KV, KSS and PTU performed the diagnostic assays and analysed the results; VV and SC performed the NGS assays; VS, AFA, RMR performed the bioinformatics analyses; VS, PTU, AFA and RMR analysed the sequencing data, PTU and VS prepared the original manuscript; VPB assisted in providing resources and reviewed the data and the final manuscript. All authors approved the final manuscript.

## Funding

The authors acknowledge the funding received from the Indian Council of Medical Research for this study, and the support from the ‘Pan India epidemiological, virological and genomic surveillance of human influenza and COVID-19 through DHR-ICMR VRDL network’ for the sequencing reagents and test kits. This work used the High Performance Computing facility “NAKSHATRA” developed under the Indian Council of Medical Research PM-Ayushman Bharat Health Infrastructure Mission (PM-ABHIM) project (Grant No. VIR/35/2022/ECD-1) for the bioinformatics analysis.

## Acknowledgements

We acknowledge Dr. Y. A. Pathan, Joint Commissioner of Animal Husbandry, Pune, and Dr. Sana Khan, WVS Hope Mission Rabies, Pune, for the case referral to our Laboratory and providing necessary details.

## Notes

### Competing Interest Statement

The authors have declared no competing interest.

### Author Declarations

The institutional review committee approved the manuscript for submission.

## References

1. World Health Organization. Rabies [Internet]. Geneva:WHO; 2024 Jun 4 [cited 2026 Jun 17]. Available from: https://www.who.int/news-room/fact-sheets/detail/rabies.

2. Swedberg C, Bote K, Gamble L, Fénelon N, King A, Wallace RM. Eliminating invisible deaths: the woeful state of global rabies data and its impact on progress towards 2030 sustainable development goals for neglected tropical diseases. Front Trop Dis. 2024 Mar;5:10.3389/fitd.2024.1303359. doi:10.3389/fitd.2024.1303359 PubMed PMID: 39811393; PubMed Central PMCID: PMC11730431.

3. Gan H, Hou X, Wang Y, Xu G, Huang Z, Zhang T, et al. Global burden of rabies in 204 countries and territories, from 1990 to 2019: results from the Global Burden of Disease Study 2019. Int J Infect Dis. 2023 Jan 1;126:136–44. doi:10.1016/j.ijid.2022.10.046

4. Troupin C, Dacheux L, Tanguy M, Sabeta C, Blanc H, Bouchier C, et al. Large-Scale Phylogenomic Analysis Reveals the Complex Evolutionary History of Rabies Virus in Multiple Carnivore Hosts. PLOS Pathog. 2016 Dec 15;12(12):e1006041. doi:10.1371/journal.ppat.1006041

5. Varun CN, Chandel S, Kumar DK, Siddalingaiah N, Telang S, Bhalke PS, et al. Comprehensive genomic analysis of rabies virus in India reveals distinct lineages and evolutionary stability. Virus Genes. 2026 Feb;62(1):92–105. doi:10.1007/s11262-025-02212-9 PubMed PMID: 41419697.

6. World Health Organization. Effective vaccination against rabies in puppies in rabies endemic regions [Internet]. Geneva: WHO; 2025 [cited 2026 Jun 17]. WHO Vaccines and Biologicals reference number: VR/102975. Available from https://www.who.int/publicatoins/i/item/vr-102975.

7. Morters MK, McNabb S, Horton DL, Fooks AR, Schoeman JP, Whay HR, et al. Effective vaccination against rabies in puppies in rabies endemic regions. Vet Rec. 2015 Aug 8;177(6):150. doi:10.1136/vr.102975 PubMed PMID: 26109286; PubMed Central PMCID: PMC4552936.

8. Madhusudana SN, Sukumaran SM. Antemortem diagnosis and prevention of human rabies. Ann Indian Acad Neurol. 2008;11(1):3–12. doi:10.4103/0972-2327.40219 PubMed PMID: 19966972; PubMed Central PMCID: PMC2781142.

9. Nadin-Davis SA, Sheen M, Wandeler AI. Development of real-time reverse transcriptase polymerase chain reaction methods for human rabies diagnosis. J Med Virol. 2009 Aug;81(8):1484–97. doi:10.1002/jmv.21547 PubMed PMID: 19551825.

10. Fastqc [Internet]. Available from: https://github.com/s-andrews/fastqc

11. Chen S, Zhou Y, Chen Y, Gu J. fastp: an ultra-fast all-in-one FASTQ preprocessor. Bioinformatics. 2018 Sep 1;34(17):i884–90. doi:10.1093/bioinformatics/bty560

12. Li H, Durbin R. Fast and accurate short read alignment with Burrows-Wheeler transform. Bioinformatics. 2009 Jul 15;25(14):1754–60. doi:10.1093/bioinformatics/btp324 PubMed PMID: 19451168; PubMed Central PMCID: PMC2705234.

13. Li H, Handsaker B, Wysoker A, Fennell T, Ruan J, Homer N, et al. The Sequence Alignment/Map format and SAMtools. Bioinformatics. 2009 Aug 15;25(16):2078–9. doi:10.1093/bioinformatics/btp352 PubMed PMID: 19505943; PubMed Central PMCID: PMC2723002.

14. Bcftools by samtools [Internet]. [cited 2026 Jul 2]. Available from: https://samtools.github.io/bcftools/

15. Seekings AH, Howard WA, Nuñéz A, Slomka MJ, Banyard AC, Hicks D, et al. The Emergence of H7N7 Highly Pathogenic Avian Influenza Virus from Low Pathogenicity Avian Influenza Virus Using an in ovo Embryo Culture Model. Viruses. 2020 Sep;12(9):920. doi:10.3390/v12090920

16. Campbell K, Gifford RJ, Singer J, Hill V, O’Toole A, Rambaut A, et al. Making genomic surveillance deliver: A lineage classification and nomenclature system to inform rabies elimination. PLOS Pathog. 2022 May 2;18(5):e1010023. doi:10.1371/journal.ppat.1010023

17. Veyseh APB, Dernoncourt F, Chang W, Nguyen TH. MadDog: A Web-based System for Acronym Identification and Disambiguation [Internet]. arXiv; 2021 [cited 2026 Jun 16]. Available from: http://arxiv.org/abs/2101.09893 doi:10.48550/arXiv.2101.09893

18. Fu L, Niu B, Zhu Z, Wu S, Li W. CD-HIT: accelerated for clustering the next-generation sequencing data. Bioinformatics. 2012 Dec 1;28(23):3150–2. doi:10.1093/bioinformatics/bts565 PubMed PMID: 23060610; PubMed Central PMCID: PMC3516142.

19. Katoh K, Standley DM. MAFFT Multiple Sequence Alignment Software Version 7: Improvements in Performance and Usability. Mol Biol Evol. 2013 Apr;30(4):772–80. doi:10.1093/molbev/mst010 PubMed PMID: 23329690; PubMed Central PMCID: PMC3603318.

20. Steenwyk JL, Iii TJB, Li Y, Shen XX, Rokas A. ClipKIT: A multiple sequence alignment trimming software for accurate phylogenomic inference. PLOS Biol. 2020 Dec 2;18(12):e3001007. doi:10.1371/journal.pbio.3001007

21. Nguyen LT, Schmidt HA, von Haeseler A, Minh BQ. IQ-TREE: A Fast and Effective Stochastic Algorithm for Estimating Maximum-Likelihood Phylogenies. Mol Biol Evol. 2015 Jan;32(1):268–74. doi:10.1093/molbev/msu300 PubMed PMID: 25371430; PubMed Central PMCID: PMC4271533.

22. Abdullah D, Susilo S, Ahmar AS, Rusli R, Hidayat R. The application of K-means clustering for province clustering in Indonesia of the risk of the COVID-19 pandemic based on COVID-19 data. Qual Quant. 2022 Jun 1;56(3):1283–91. doi:10.1007/s11135-021-01176-w

23. Kosakovsky Pond SL, Frost SDW. Not So Different After All: A Comparison of Methods for Detecting Amino Acid Sites Under Selection. Mol Biol Evol. 2005 May 1;22(5):1208–22. doi:10.1093/molbev/msi105

24. Murrell B, Wertheim JO, Moola S, Weighill T, Scheffler K, Pond SLK. Detecting Individual Sites Subject to Episodic Diversifying Selection. PLOS Genet. 2012 Jul 12;8(7):e1002764. doi:10.1371/journal.pgen.1002764

25. Weaver S, Shank SD, Spielman SJ, Li M, Muse SV, Kosakovsky Pond SL. Datamonkey 2.0: A Modern Web Application for Characterizing Selective and Other Evolutionary Processes. Mol Biol Evol. 2018 Mar 1;35(3):773–7. doi:10.1093/molbev/msx335

26. Maurice NA, Luka PD, Maurice MN, Ngbede EO, Zhakom PN, Mshelbwala PP, et al. RABIES IN A SET OF EIGHT-WEEK OLD PUPPIES IN NIGERIA: THE NEED FOR REVIEW OF CURRENT DOG ANTIRABIES VACCINATION SCHEDULE. Afr J Infect Dis. 2018 Jun 18;12(2):72–7. doi:10.21010/ajid.v12i2.12 PubMed PMID: 30109290; PubMed Central PMCID: PMC6085740.

27. Singhai M, Jaiswal R, Siddiqui C, Tiwari S, Gupta N, Bala M, et al. Rabies can be a disease of puppyhood. J Fam Med Prim Care. 2022 Jun;11(6):3339–41. doi:10.4103/jfmpc.jfmpc_1605_21 PubMed PMID: 36119256; PubMed Central PMCID: PMC9480633.

28. Brunker K, Jaswant G, Thumbi SM, Lushasi K, Lugelo A, Czupryna AM, et al. Rapid in-country sequencing of whole virus genomes to inform rabies elimination programmes. Wellcome Open Res. 2020;5:3. doi:10.12688/wellcomeopenres.15518.2 PubMed PMID: 32090172; PubMed Central PMCID: PMC7001756.

29. Fisher CR, Streicker DG, Schnell MJ. The spread and evolution of rabies virus: conquering new frontiers. Nat Rev Microbiol. 2018 Apr;16(4):241–55. doi:10.1038/nrmicro.2018.11 PubMed PMID: 29479072; PubMed Central PMCID: PMC6899062.

30. Conselheiro JA, Barone GT, Miyagi SAT, de Souza Silva SO, Agostinho WC, Aguiar J, et al. Evolution of Rabies Virus Isolates: Virulence Signatures and Effects of Modulation by Neutralizing Antibodies. Pathogens. 2022 Dec 19;11(12):1556. doi:10.3390/pathogens11121556 PubMed PMID: 36558890; PubMed Central PMCID: PMC9782306.

31. Riedel C, Hennrich AA, Conzelmann KK. Components and Architecture of the Rhabdovirus Ribonucleoprotein Complex. Viruses. 2020 Aug 29;12(9):959. doi:10.3390/v12090959 PubMed PMID: 32872471; PubMed Central PMCID: PMC7552012.

